# Impact of residual intimal flap displacement post-TEVAR on TBAD haemodynamics in compliant, patient-specific CFD simulations informed by MRI

**DOI:** 10.1101/2024.09.25.24313546

**Authors:** Louis Girardin, Niklas Lind, Hendrik von Tengg-Kobligk, Stavroula Balabani, Vanessa Díaz-Zuccarini

## Abstract

We propose a novel formulation of a moving boundary method to account for the motion of the intimal flap (IF) in a TBAD post-thoracic endovascular aortic repair using patient-specific compliant computational fluid dynamics simulations. The simulations were informed by non-invasive 4DMRI sequences. Predicted flow waveforms, aortic wall and IF displacements were validated against in vivo 4DMRI and cine MRI data. The patient-specific simulation showed that at peak systole, the dynamic interplay between high IF displacement and high transmural pressures promoted true lumen compression and false lumen expansion, while luminal patterns were reversed at the deceleration phase. High vorticity and swirling flow patterns were observed throughout the cardiac cycle at the primary entry tear, the descending aorta and proximal to the visceral aortic branches, correlating with high relative residence time, which could indicate an increased localised risk of aortic growth proximal to the IF. A rigid IF simulation revealed significant discrepancies in haemodynamic metrics, highlighting the potential mispredictions when using a rigid wall assumption to assess disease progression. Simulations assuming a more compliant IF highlighted increased risks of visceral branches malperfusion and localised aortic wall degeneration. The study underscores the necessity of patient-specific-compliant IF simulations for accurate TBAD hemodynamic assessments. These insights can improve disease understanding and inform future treatment strategies.

## Introduction

Type-B aortic dissection (TBAD) represents a life-threatening cardiovascular condition requiring prompt diagnosis and management to mitigate high mortality and morbidity risks. TBAD involves a primary entry tear (PET) at the descending aorta (DA), where blood seeps in the layers of the true lumen (TL), which creates the false lumen (FL); lumina are separated by the intimal flap (IF) [1]. The dissection can spread along the aorta and its branches, causing abnormal blood flow, high pressures and organ malperfusion, ultimately leading to potentially adverse outcomes. With an annual incidence affecting 1.6 in 100,000 individuals, TBAD requires close monitoring and patient-specific intervention planning to improve patient survival [2].

Surgical interventions for TBAD include thoracic endovascular aortic repair (TEVAR) and open surgery. TEVAR targets the coverage of the FL to restore normal aortic function. However, the geometric complexity of TBAD may pose challenges in achieving complete coverage, particularly when the dissection extends into branches or distally [3]. Open surgery aims to seal the PET and promote blood flow into the TL to alleviate malperfusion. Extensive surgical replacement of the affected portion should be avoided to minimise coverage of collateral arteries and prevent prioritisation of lengthy synthetic grafts, which reduce aortic compliance [4], [5]. Despite these approaches, both surgical techniques can result in a residual dissection involving a mobile IF, complicating the restoration of optimal aortic function [6].

While anatomical markers are commonly used to assess disease progression, evaluating the IF movement and its impact on transmural pressure (TMP) can aid in predicting disease progression [7], [8]. The IF influences the pressure in the lumina and determines which pressure (either in the TL or FL) is dominant at any point in the cardiac cycle. There is a complex interplay between these pressure fluctuations, blood flow volume changes and the expansion and contraction of the lumina, which has been reported to promote growth and disturbed hemodynamics [9], [10].

Increased aortic pressures are correlated with aortic wall thickening and rigidification, leading to increased heart load [11], [12]. Moreover, pressure reflections can disrupt flow dynamics and alter wall shear stress, potentially contributing to aneurysmal growth and thrombotic events [13], [14].

4D flow MRI (4DMRI) is an advanced imaging technique that has revolutionised the visualisation and quantification of complex flow patterns within the cardiovascular system, particularly in TBAD [15], [16]. However, several challenges still need to be addressed for widespread clinical implementation; 4DMRI acquisitions are costly and only available in some clinics [17]. Additionally, the large data volumes necessitate compromises in spatial and temporal resolution, typically 1-5mm and 35-50ms for aortic imaging [18], which limits the transient measurement of IF movement [19], [20]. Computational fluid dynamics (CFD) simulations can enhance these imaging modalities, providing a deeper understanding of the impact of IF movement on aortic hemodynamics and estimate parameters that cannot be measured directly [21]. Nevertheless, many CFD studies still rely on literature-based parameters as high-quality datasets are complex to obtain.

Compliant models are essential for investigating the influence of IF movement on TBAD haemodynamics, with fluid-structure interaction (FSI) being commonly employed. A recent FSI study suggested that PET size can substantially affect the TMP and haemodynamic parameters in TBAD, potentially impacting disease progression [22]. Using an idealised TBAD geometry with a constant IF thickness, [23], investigated the effect of IF motion on flow in acute TBAD using FSI. They found that IF motion increased flow into the FL and predicted higher pressures than rigid wall models. FSI simulations correlated near-PET haemodynamics and IF motion with potential thrombus formation [24]. FSI was used to demonstrate that patient-specific IF displacements must be simulated for accurate cyclical deformation and good agreement against *in vivo* 4DMRI [25]. It was also reported that the complexity of accurately measuring local IF stiffness and thickness could significantly impact the accuracy of simulations.

While being able to provide insight on the impact of the wall and IF on TBAD haemodynamics, FSI-based studies often acknowledge the lack of *in vivo* tissue data to describe patient-specific material properties in the aortic wall and the IF [26]. To overcome this limitation, we have developed a moving boundary method (MBM), an alternative to FSI that uses a deformable mesh and does not require a detailed structural model of the arterial wall to capture the interactions between the fluid and the wall. The MBM is informed by patient-specific clinical images and results in substantial gains in computational time [27]. In this method, the displacement of the aortic external wall and IF follows the local surface-normal direction. It is linearly related to the fluid forces acting on them, with stiffness coefficients tuned based on patient-specific displacement data. However, in our previous work, the IF was modelled as a zero-thickness membrane with small displacements, which limits the accuracy of representing cross-sectional area variations [28].

In this paper, we analyse the impact of patient-specific IF displacements on the haemodynamics of a TBAD case post-TEVAR with a remaining IF with varying thickness via an improved MBM. Following our previous work [29], 4DMRI data are exclusively utilised to inform a patient-specific-compliant CFD simulation, which increases the accuracy and relevance of the findings for the individual patient, and the predictions are validated against brachial pressures, 4DMRI and cine-MRI. Additional simulations, including a rigid IF simulation and two others in which the IF exhibits lower degrees of stiffness leading to higher displacement are compared against the patient specific case. Pressure and velocity magnitude distributions, the extent of the rotational flow, TMP, and wall shear stress-indices (WSS) are employed to characterise the impact of IF displacement on hemodynamics.

## 1. Materials and Methods

### 1.1. Clinical Data

A patient with chronic type B aortic dissection (TBAD) previously treated with thoracic endovascular aortic repair (TEVAR) underwent follow-up imaging at Inselspital Bern, in accordance with an ethically approved protocol. This study was reviewed and approved by the Cantonal Ethics Committee of Bern (Kantonale Ethikkommission für die Forschung), under the jurisdiction of the Gesundheits-und Fürsorgedirektion des Kantons Bern (Health and Welfare Directorate of the Canton of Bern). The approval reference number for this study is 2019-00556, and the decision of the comitee was issued on July 30, 2019.Brachial pressures were measured before the imaging procedures. MRI sequences were performed on a MAGNETOM Sola fit scanner (Siemens Healthineers). The imaging protocol included acquiring 4DMRI, two planes of cine-MRI, and T2/T1 weighted TRUFI MRI sequences, with respective pixel sizes of 2.5*2.5*2.5 mm^3^ 1.88*1.88 mm^2^ and 1*1*1 mm^3^, to visualise the patient’s thoracic aorta.

### 1.2. Segmentation and Meshing

The aortic geometry was obtained from the T2/T1 TRUFI utilising semi-automatic segmentation and smoothing techniques implemented in ScanIP (Synopsis Simpleware, USA). The geometry preparation and meshing were performed using Fluent Mesh (Ansys Fluent, USA). The IF was subdivided into pairs of patches (with each patch facing the TL and FL respectively) of approximately 10 mm along the centerline, following the curvature of the IF. This subdivision was designed to ensure that within each patch pair, the surface normals of all nodes are sufficiently similar. This approximation allows smooth nodal displacement within each patch pair, facilitating the application of the MBM (Fig 1c). Specifics regarding mesh element sizing, parameters applied for prism layering, and the mesh sensitivity analysis conducted to achieve the final mesh are provided in the Appendix.

**Fig 1.**
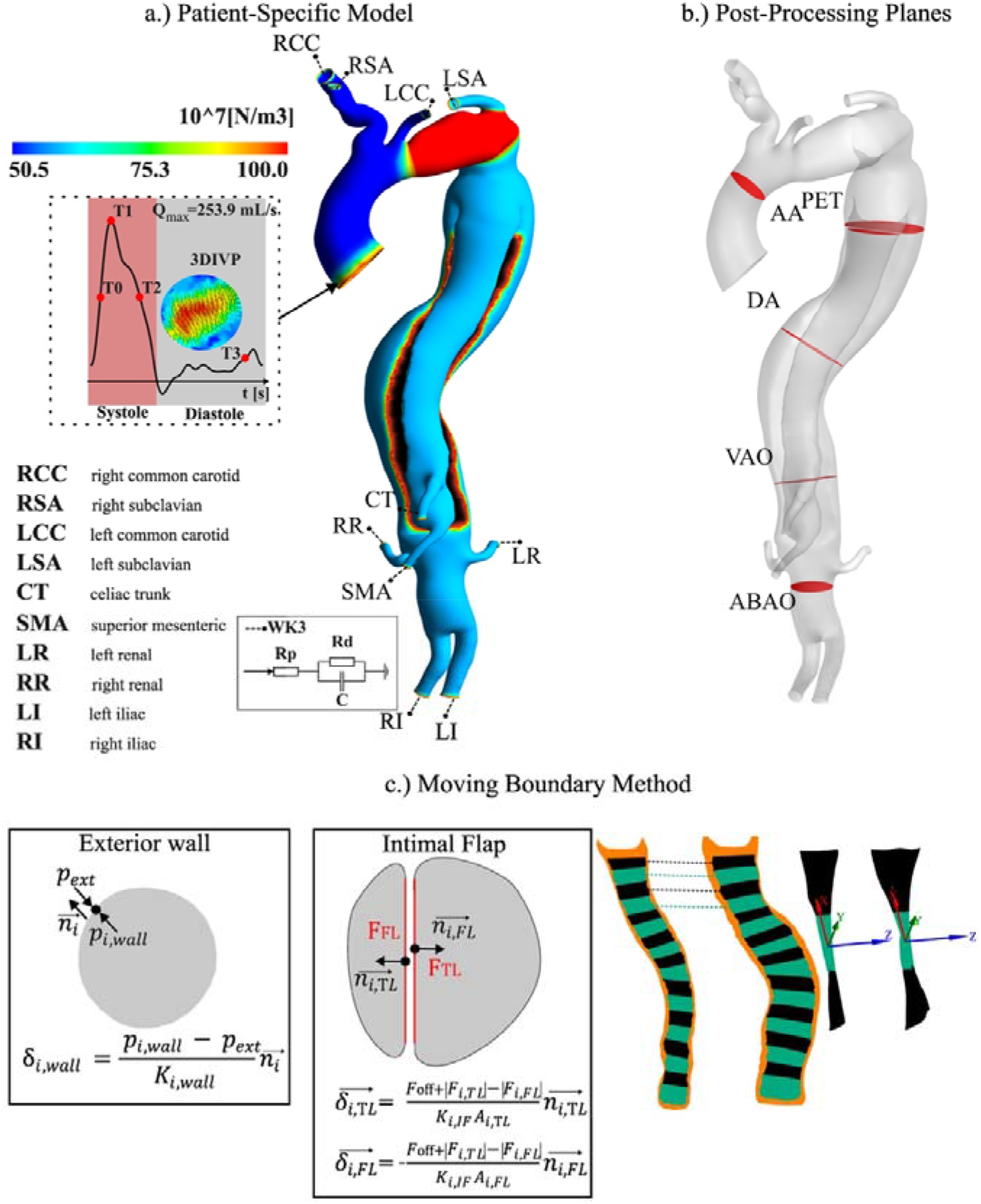
a.) Patient-specific model of the domain showing the boundary conditions used and the aortic wall stiffness. T0 is mid-acceleration, T1 is peak systole, T2 is mid-deceleration, and T3 is end of diastole. b.) Planes in red are used to extract variables in the post-processing at the ascending aorta (AA), PET, descending aorta (DA), visceral aorta (VAO) and abdominal aorta (ABAO). c.) The MBM is applied both at the exterior aortic wall and the IF. The rightmost part of the figure illustrates the patching technique used for the IF, showing how TL/FL pairs of patches, coloured in black and green respectively, share the same surface normal.

### 1.3. Moving Boundary Method (MBM)

The MBM previously proposed by Bonfanti et al., 2017, and compared against FSI [28] was further developed so that it could be applied to simulate the displacement of the entire aorta, i.e., the aortic wall and the thick IF.

#### 1.3.1. Aortic Wall

The local exterior wall displacement *δ*_*i,wall*_ (m) is defined as:

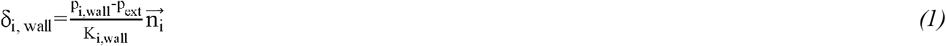

Where 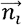 is the local unit normal vector, p_i,wall_ (Pa) the aortic wall nodal pressure and p_ext_ (Pa), the exterior pressure set as the diastolic pressure, and K_i,wall_ (N/m^3^) is the exterior, local wall stiffness. The stiffness is derived from the distensibility, estimated from the 4DMRI data and defined as the ratio between the crosssectional relative change and the regional pulse pressure (Figure 1a; see previous works by Bonfanti et al., 2017, for a detailed method description).

#### 1.3.2. Intimal Flap

As described in section 1.2, the TL and FL sections of the IF were discretised into patches along the centerline (Figure 1c). Each patch in the TL section was paired with the nearest patch in the FL section on the opposite side of the IF. This pairing ensured that the displacement of facing IF patches is synchronised, thereby preserving the thickness of the IF.

The displacement of a pair of patches is proportional to the normal force gradient of the patches and inversely proportional to a local stiffness coefficient K_i,IF_ (N/m^3^) along the surface normal, such as:

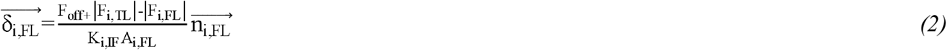

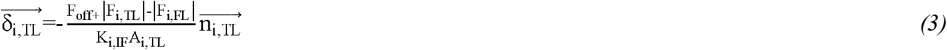

Where TL and FL denote true and false lumen, respectively, δ_i,TL_ and δ_i,FL_ (m) are the displacement, F_i,TL_ and F_i,FL_ (N) the average forces, A_i,TL_ and A_i,FL_ (m^2^) the surface areas, 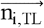 and 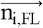 the surface normal of each respective *i* patch TL/FL pair. F_off_ (N) is the pre-stress force measured on the rigid IF simulation used to start the displacement from zero and to avoid a displacement ‘jump’ at the first simulation time step. K_i,IF_ was iteratively tuned to match the patient-specific displacement measured on the cine-MRI.

Four IF stiffness values were then considered: a rigid one (named D0), the patient-specific stiffness (D1) derived from clinical images, and two additional cases where the stiffness is two times smaller (D2) and three times smaller (D3) than in the patient-specific case. As shown in previous work, a smoothing algorithm was used between regions of different stiffness to avoid abrupt displacement transitions [30].

### 1.4. Inlet and Outlet Boundary Conditions

Following our previous work [31], 4DMRI was used to extract the three-dimensional inlet velocity profile (3DIVP) (Figure 1a) and outlet mean flow rates using GTFlow (GyroTools LLC, Switzerland) (Table 1). MATLAB (MathWorks Inc., USA) was used to spline-interpolate the inlet flow rate to apply a 1 ms time-step for the CFD simulations.

**Table 1.** Targeted values of inlet pressures and outlet mean flow rates against simulation results.

|  |  | Target | D0 | D1 | D2 | D3 |
| --- | --- | --- | --- | --- | --- | --- |
| **Pressure**<br>**(mmHg)** | **Diastole** | 84.00 | 82.9/1.3% | 82.8/1.5% | 82.5/1.7% | 82.1/2.3% |
|  | **Systole** | 113.86 | 113/0.7% | 114/-0.1% | 114.9/-0.9% | 115.6/-1.5% |
| **Mean Flow**<br>**Rate**<br>**(mL/s)** | **RCC** | 10.02 | 10/-0.2% | 10/0.1% | 10/0.0% | 10/-0.2% |
|  | **RSA** | 6.82 | 6.8/-0.3% | 6.8/-0.7% | 6.9/-3.7% | 7.1/-1.5% |
|  | **LCC** | 6.55 | 6.5/-0.3% | 6.6/0.1% | 6.5/0.4% | 6.5/0.1% |
|  | **LSA** | 10.42 | 10.4/1.1% | 10.3/0.6% | 10.4/0.3% | 10.4/0.7% |
|  | **CT** | 3.51 | 3.5/2.3% | 3.4/-1.4% | 3.6/0.6% | 3.5/1.5% |
|  | **SMA** | 5.72 | 5.7/-2.1% | 5.8/-1.7% | 5.8/-2.1% | 5.8/-4.5% |
|  | **LR** | 6.83 | 6.8/-0.4% | 6.9/0.6% | 6.8/3.3% | 6.6/3.3% |
|  | **RR** | 8.35 | 8.4/2.8% | 8.1/1.2% | 8.3/2.7% | 8.1/2.1% |
|  | **LI** | 6.28 | 6.3/-5.2% | 6.6/-1.2% | 6.4/-4.8% | 6.6/-4.6% |
|  | **RI** | 5.57 | 5.6/2.0% | 5.5/0.9% | 5.5/2.5% | 5.4/2.5% |

A zero-dimensional lumped parameter model of the aorta was built and tuned in 20-sim (Controllab Products, Netherlands), targeting the *in vivo* inlet pressures and outlet mean flow rates (Table 1). Three-element Windkessel (WK3) pressure conditions were used at the domain outlets as described in past works (Figure 1a) [32], [33]. The WK3 parameters are provided in the Appendix.

### 1.5. Computational Model

The finite-volume solver ANSYS CFX 2023R2 was utilised to solve the transient three-dimensional Navier-Stokes equations, modelling blood as an incompressible and non-Newtonian fluid with a density of 1056 kg/m^3^ and viscosity described by the Carreau-Yasuda model with empirical constants given by Tomaiuolo et al., 2016. The peak Re_p_ and critical Re_c_ Reynolds numbers were calculated as 8262 and 7407, respectively [35], indicating turbulent flow conditions, so the k-ω shear stress transport model was employed to capture turbulence effects. A low turbulence intensity of 1% was introduced to account for the laminar-turbulent transition [36]. The numerical simulations were conducted with a second-order backward Euler scheme, with convergence criteria set to a root-mean-square residual value of 10^−5^ for all equations within each time step. Periodic behaviour characterised by less than 1% variation in systolic and diastolic pressures between cycles was achieved after four cycles for all simulations after appropriate initialisation. The results from the final cycle were post-processed to extract relevant hemodynamic indices.

### 1.6. Haemodynamics Analysis

To assess the impact of IF displacement on the TL and FL haemodynamics, TMP, vorticity, in-plane rotational flow (IRF) and WSS-driven metrics were estimated and compared for all the cases simulated.

TMP (mmHg) is the pressure difference between TL and FL:

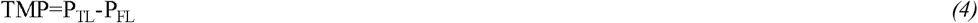

TMP values were extracted along the IF centerline, every 20 mm from the PET when possible for four points in the cardiac cycle: mid-acceleration (T0), peak systole (T1), mid-deceleration (T2), and end of diastole (T3).

Vorticity (1/s) was used to visualise the rotational characteristics of the blood flow. The component orthogonal to the cross-sectional planes of the aortic geometry (Figure 1b) was calculated as follows:

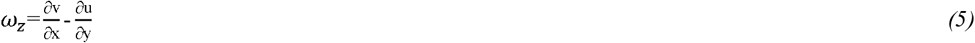

This vorticity was then integrated to produce the in-plane rotational flow (IRF) (m^2^/s) metric, which quantifies the strength of vorticity and has been correlated to the expansion of the FL [37]:

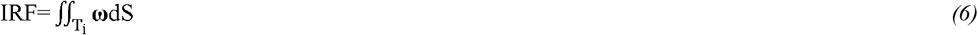

In TBAD with residual and mobile IF, IRF measurements can provide insight into the altered haemodynamic environment and highlight potential areas contributing to the growth of the FL. The in-plane rotational flow was calculated at four points in the cardiac cycle similar to TMP, namely T0, T1, T2 and T3 at the following locations: AA, PET, DA, VAO and ABAO (see Figure 1b).

Time averaged wall shear stress (TAWSS) (Pa), oscillatory shear index (OSI) and relative residence time (RRT) (Pa^-1^) were calculated every 5 ms [38]. These metrics are typically used to provide insights into the complex flow dynamics and potential risk areas in the aorta. Regions of low TAWSS, high OSI, and elevated RRT have been associated with adverse remodelling and potential complications in TBAD patients [39]. These metrics were computed as follows:

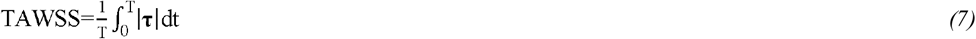

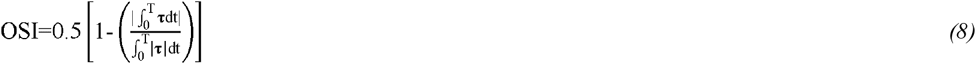

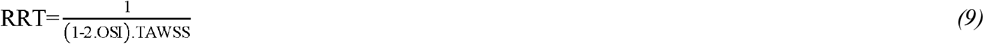

where T is the cardiac cycle period (s), and τ the instantaneous WSS.

## 2. Results

### 2.1. Validation

#### 2.1.1. Pressure and Flows

In the patient-specific case (D1, Table 1), target mean flow rates and pressures are simulated with maximum errors of 1.7% and 1.5%, respectively. Aortic pressures are closely matched in all other simulations. However, in D2 and D3, where the flap is more mobile, mean flow rates are simulated with errors up to 4.5% at the visceral branch outlets.

A good qualitative agreement is observed between D1 and the 4DMRI measurements (Figure 2). The velocity magnitude distributions at T0, T1, and T2 are well captured, especially at the PET, DA, and VAO, where the patient-specific IF displacement impacts the flow. The 3DIVP shows a good agreement with 4DMRI at the AA in both cases. A low 4DMRI signal-to-noise ratio at the end of diastole (T3) likely explains the poor agreement between D1 and 4DMRI velocity distributions. Discrepancies are observed proximal to the IF in the rigid flap simulation (D0) compared to D1 and 4DMRI velocities.

**Fig 2.**
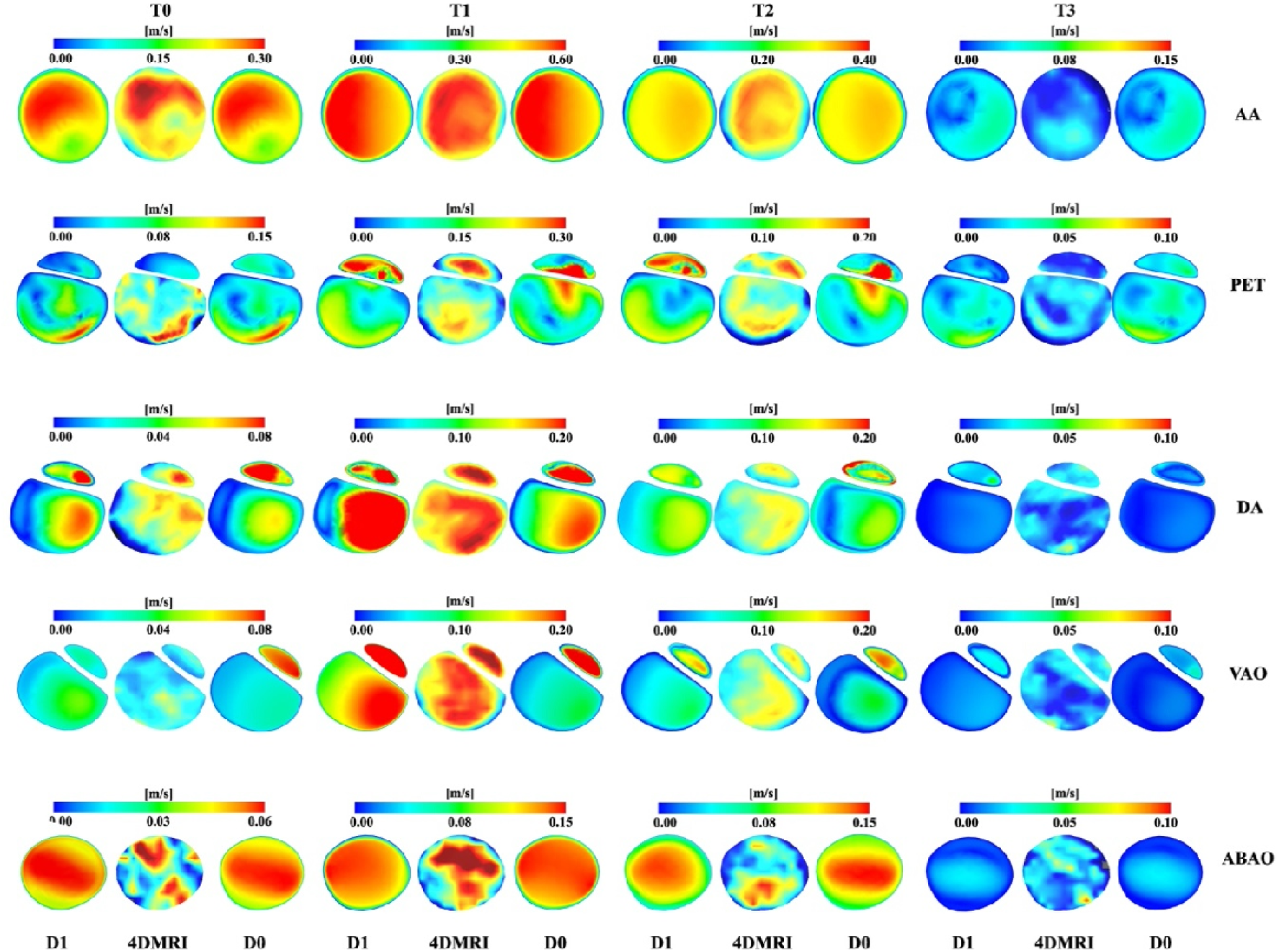
Velocity magnitude comparisons between patient specific simulations (D1), 4DMRI and rigid wall simulations (D0) at mid-acceleration (T0), peak systole (T1), mid-deceleration (T2), and end of diastole (T3) in selected locations (AA, PET, DA, VAO and ABAO).

Notable mispredictions occur in the location of the peak velocity magnitude, the size of the distribution of these peak values, and the peak values themselves at the PET, DA, and VAO across T0, T1, and T2. At T0, both simulations agree with the 4DMRI. However, only D1 corresponds well with the 4DMRI at the DA and VAO, while D0 shows an excessively high FL velocity at the VAO. At T1, D1 captures the high-velocity regions at both the PET and DA, closely matching the 4DMRI, whereas D0 presents lower and inaccurate velocity distributions. At the VAO, D1 mimics the 4DMRI pattern, but the TL velocity distribution in D0 is poorly captured. At T2, D1 aligns well with the 4DMRI, particularly at the TL at the PET and DA. However, D0 exhibits lower velocities and incorrect distribution patterns, especially at both lumina.

#### 2.1.2. Patient-Specific Wall and IF Displacements

Cine-MRI is used to validate the model by comparing the predicted displacements of the patient-specific case (D1) at the AA and VAO (Figure 3). The resolution of the cine-MRI does not allow for the measurement of transient displacements of the aortic wall and IF; only the peak deformations at T1 and the diastolic (T3) cross-section are measured and compared. The patient-specific simulations (D1) capture both the magnitude and pattern of the wall displacement between T1 and T3, with TL being compressed and FL expanding at T1.

**Fig 3.**
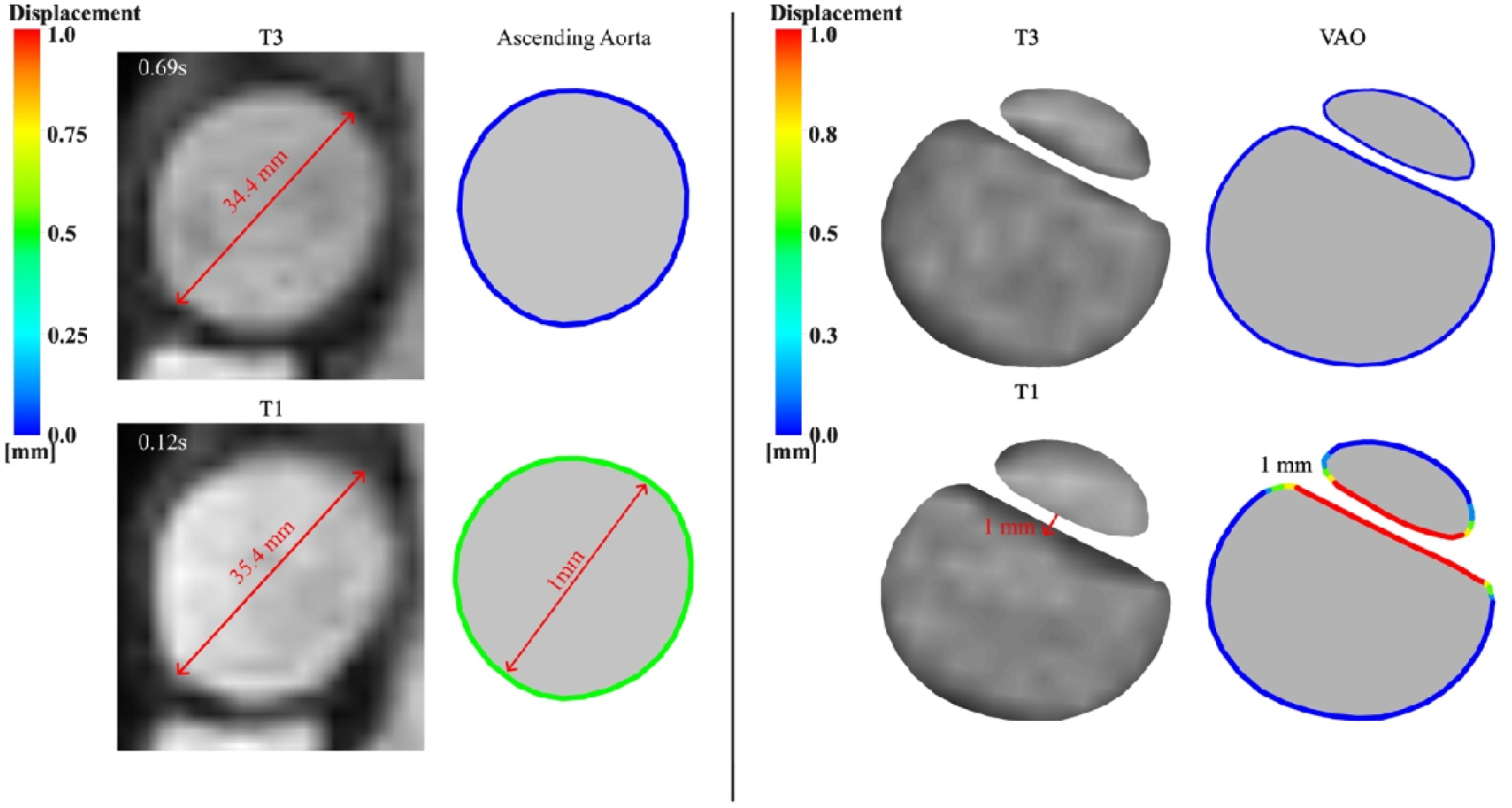
Validation of the aortic wall and IF displacement in the patient-specific case D1 against cine-MRI measurements. The left panel in each subset shows the cine MRI and the right one shows the simulated crosssections, with the boundary colour indicating the magnitude of the displacement. A red arrow highlights the IF displacement.

### 2.2. IF Displacement, TMP and Pressure Contours

Figure 4 illustrates the displacement of the IF at the PET, DA, VAO, and TMP along the IF for each case at different points in the cardiac cycle (T0, T1, T2, and T3). In D1, the maximum IF displacement reaches 0.5, 0.6, and 1 mm at the PET, DA, and VAO, respectively. Similarly, TMP values increase with the distance from the PET at each time point. Specifically, the TMP is negative at T0, T2, and T3 and increases linearly, with a maximum of about -4 mmHg. At T2, the TMP is positive and correlates with the decreasing part of the displacement curve when the IF moves back towards its diastolic position and the FL is compressed.

**Fig 4.**
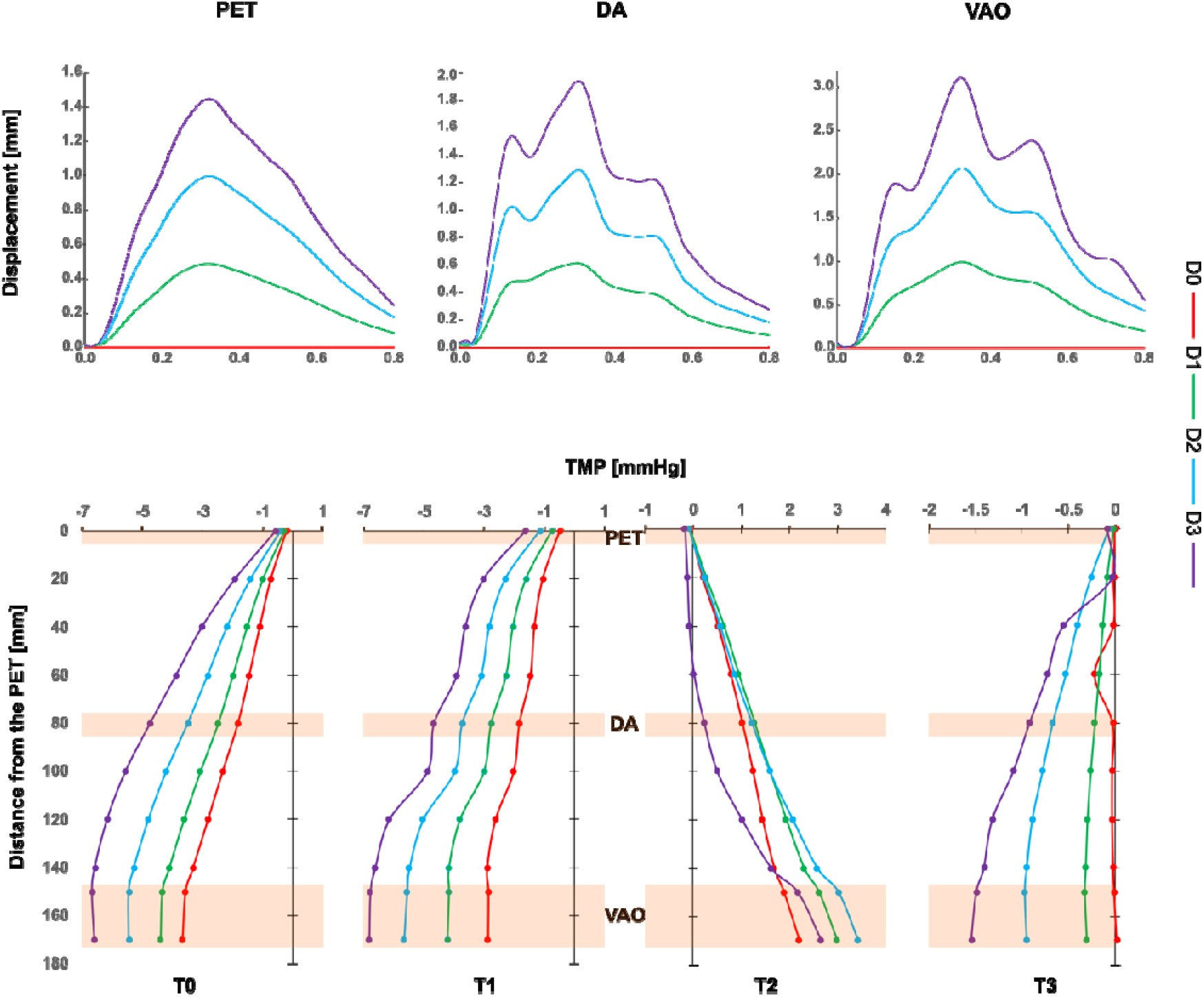
IF displacement at the PET, DA and VAO over the cardiac cycle and TMP starting from the PET at T0, T1, T2 and T3 for all cases.

In the rigid simulations (D0), the IF does not move. The TMP follows similar trends at the T0, T1, and T2 time points, although the values are lower. However, at diastole (T3), the TMP remains close to zero, with a slight TMP increase proximal to the DA, which does not compare well with D1.

The simulations with more compliant IF, D2 and D3, follow similar trends at T0 and T1 compared to the patient-specific case D1, with higher TMP values correlating with higher IF displacements, up to -7 mmHg for D3. However, these trends are not replicated at T2 and T3. Specifically, all TMP values in D3 are smaller than those of D1 at T2, and D2 and D1 curves are similar in proximal locations down to 120 mm. Finally, at T3, the TMP curve proximal to the PET in D3 does not compare well with D1, as a sharp increase is observed.

The pressure distribution for the patient specific case (D1) is shown in Figure 5 for two instants in the cardiac cycle (T1 and T2). The FL is highly pressurized at T1, with a pressure of 116 mmHg. Conversely, at T2, the FL becomes more pressurized and compressed. Comparisons against the pressure values obtained in the other cases indicate lower pressures for D0, especially at T1 in the FL, where a maximum difference of 2.4 mmHg is observed. On the contrary, the D2 and D3 cases exhibit higher pressures, reaching up to 122.34 mmHg at the visceral branches in D3, close to the location of the highest IF displacement.

**Fig 5.**
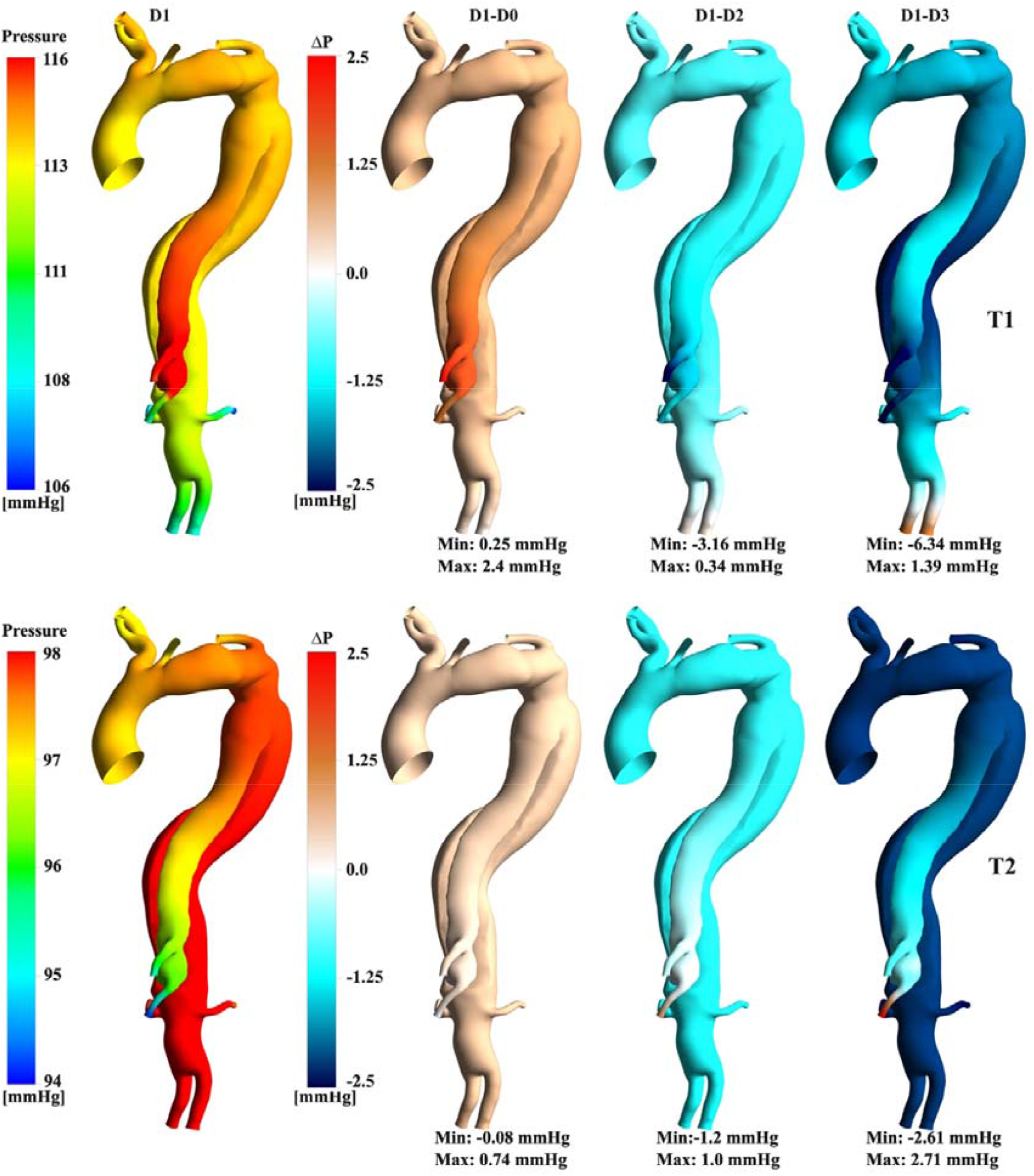
Pressure contours for D1 and point-wise difference against additional cases at T1 and T2. Maximum and minimum point-wise differences are reported below each comparison.

### 2.3. Area Change Over the Cardiac Cycle

The cross-sectional area variation over the cardiac cycle is plotted at multiple locations to demonstrate the compliant behavior of each case (Figure 6). As expected, the displacement at AA and ABAO is the highest. Additionally, the interplay between FL expansion and TL contraction is depicted at the IF locations, with the maximum FL expansion occurring at the VAO for D3, reaching about 25%.

**Figure 6.**
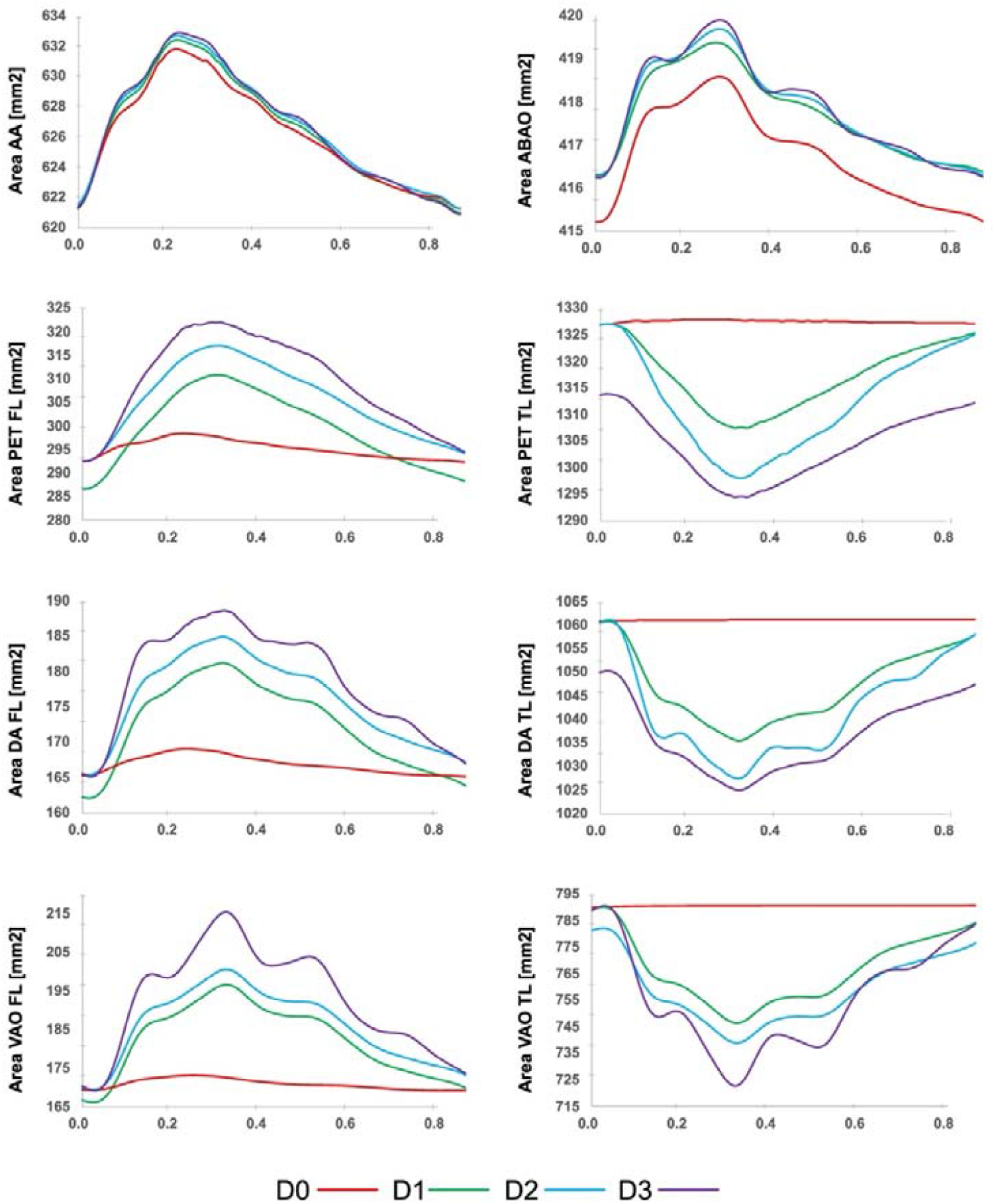
Cross-sectional area variation over the cardiac cycle in selected locations for all simulated cases.

### 2.1. Rotational Flow Features

Vorticity patterns are analyzed at selected locations (PET, DA, and VAO). In D1, vorticity patterns at PET reveal ongoing dynamic interactions between the TL and FL. In the PET region, counter-rotating vortices are clearly present in the TL, with vorticity peaking at peak systole (T2). This indicates significant rotational flow dynamics that may influence the motion of the flap separating the TL and FL. The persistent vorticity in the FL near the PET, even after TEVAR, suggests the presence of non-uniform flow that could contribute to flap displacement, potentially impacting FL expansion. In the DA region, vorticity is less pronounced, indicating lower recirculation and a more uniform flow pattern compared to the PET region. This suggests that the flow dynamics in the DA are less likely to significantly influence flap motion. However, in the VAO region, despite overall lower vorticity values, some circulatory patterns are still observed in T1. These circulations, although weaker, might still affect the flow dynamics, potentially influencing the hemodynamic environment near the VAO.

Comparing the patient specific (D1) case against the rigid wall (D0) one, differences in vorticity patterns can clearly be seen highlighting the impact of IF motion on flow dynamics. Distinct vorticity values, strengths, and swirling patterns are observed at the PET and the VAO, especially during systole (T0, T1 and T2), which suggest changes in WSS. As the IF becomes more mobile, vorticity extrema are globally enhanced in D2 and D3 compared to D1.

The rotational flow characteristics observed across different regions, particularly at the FL, are reflected in the IRF values summarised in Table 2, demonstrating how differences in IF treatment influence the hemodynamic environment. In D1, IRF values tend to increase from T0 to T1 and then decrease during the deceleration phases, regardless of the sign of the value, at the AA, DA, VAO, and ABAO (Table 2). High IRF magnitudes (>20 cm^2^/s) are particularly noted in the AA, PET, and VAO at T1, suggesting significant rotational flow in these regions. Conversely, IRF values at the DA remain close to zero, indicating a balance between positive and negative vorticity. In D0, significant differences are simulated compared to D1, particularly at the PET. These disparities are most pronounced at T1, where the IRF is underpredicted in the TL and overpredicted in the FL, impacting the accuracy of growth predictions proximal to the PET. Additionally, the magnitude of the IRF suggests an increased risk of growth at the PET in simulations with more mobile intimal flap displacement (D2 and D3).

**Table 2.** IRF measured at the AA and ABAO, and in both lumina at the PET, DA, and VAO for every case at T0, T1, T2 and T3.

|  |  | IRF[cm<sup>2</sup>/s] |  |  |  |  |  |  |  |
| --- | --- | --- | --- | --- | --- | --- | --- | --- | --- |
|  |  | D0 |  | D1 |  | D2 |  | D3 |  |
|  |  | T0 | T1 | T0 | T1 | T0 | T1 | T0 | T1 |
| AA |  | 5.51 | 34.30 | 5.54 | 34.22 | 5.60 | 34.08 | 5.98 | 33.92 |
| PET | TL | 5.13 | 25.72 | 5.30 | 33.92 | 5.47 | 44.56 | 5.63 | 56.37 |
|  | FL | -10.65 | -13.33 | -11.34 | -6.01 | -12.71 | 2.21 | -13.87 | 11.20 |
| DA | TL | -0.09 | -1.11 | -0.11 | -0.99 | -0.14 | -0.83 | -0.18 | -0.61 |
|  | FL | 0.29 | -1.84 | 0.21 | -1.48 | 0.15 | -1.05 | 0.02 | -0.57 |
| VAO | TL | -2.84 | -19.47 | -3.53 | -18.15 | -4.97 | -15.86 | -6.83 | -12.47 |
|  | FL | -0.23 | -0.7 | -0.18 | -0.52 | -0.13 | -0.27 | -0.03 | 0.059 |
| ABAO |  | 0.66 | 5.38 | 0.51 | 5.62 | 0.44 | 5.95 | 0.33 | 6.36 |
|  |  | T2 | T3 | T2 | T3 | T2 | T3 | T2 | T3 |
| AA |  | 22.94 | 1.22 | 22.9 | 1.12 | 22.86 | 0.95 | 22.79 | 0.78 |
| PET | TL | 50.69 | -4.5 | 53.24 | -3.53 | 57.25 | -3.48 | 61.23 | -2.82 |
|  | FL | 21.50 | -7.18 | 25.43 | -8.62 | 30.16 | -8.77 | 36.57 | -4.25 |
| DA | TL | -0.72 | -0.11 | -0.51 | -0.23 | -0.34 | -0.45 | -0.09 | -0.89 |
|  | FL | -0.26 | -0.07 | -0.18 | -0.13 | -0.13 | -0.19 | -0.01 | -0.24 |
| VAO | TL | -0.73 | 0.12 | -0.81 | 0.65 | -0.89 | 1.28 | -1.11 | 2.12 |
|  | FL | -0.26 | -0.34 | -0.63 | -0.33 | -0.99 | -0.37 | -1.34 | -0.43 |
| ABAO |  | 3.89 | 0.85 | 4.01 | 0.84 | 4.3 | 0.84 | 4.78 | 0.74 |

### 2.2. Wall Shear Stress Indices

Figures 8-10 display the contours of time-average wall shear stress (TAWSS), oscillatory shear index (OSI), and relative residence time (RRT) obtained for the patient-specific case (D1), alongside point-wise differences of these metrics from the additional cases.

**Figure 7.**
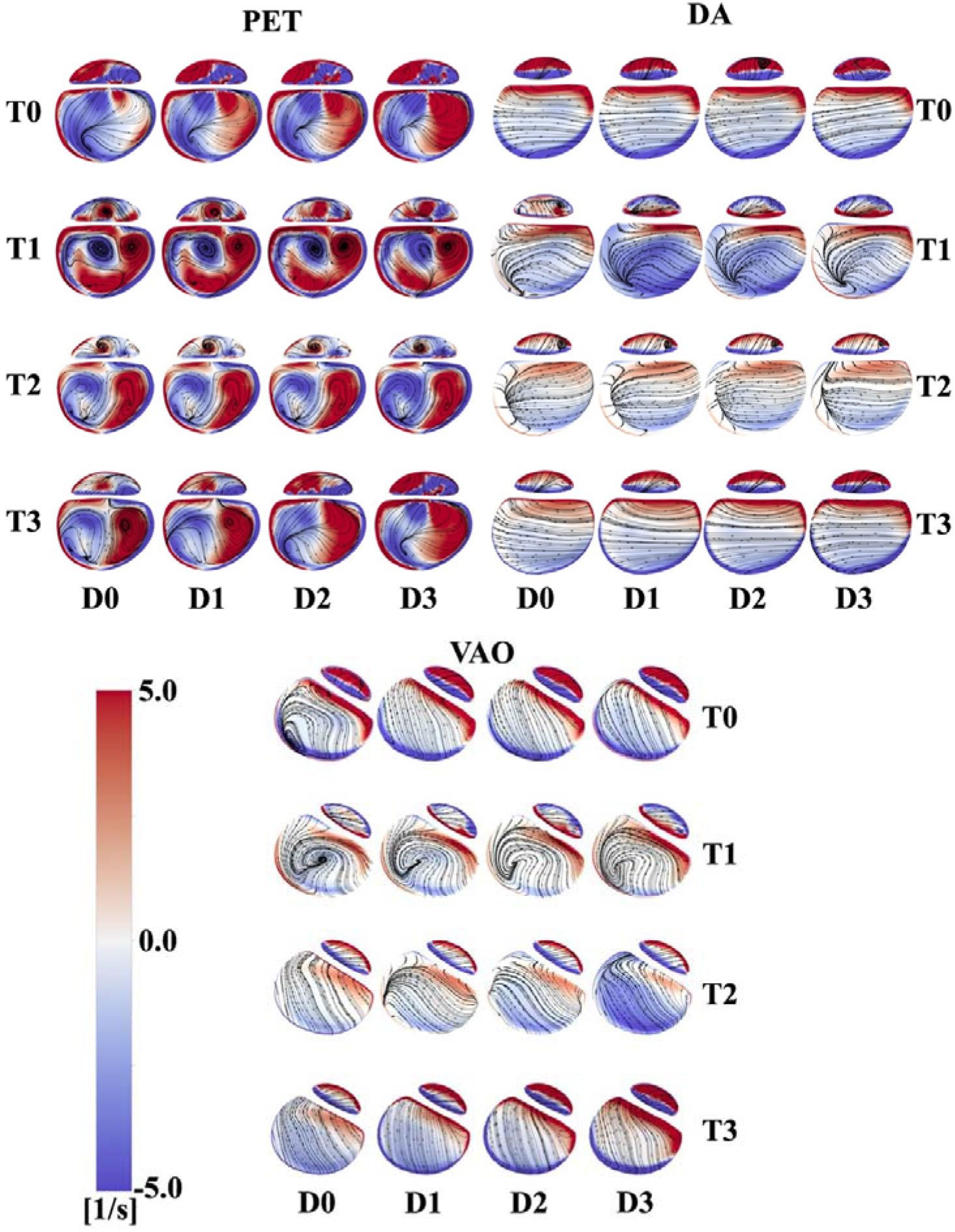
Vorticity contours and overlapping streamlines for every case at the PET, DA and VAO at T0, T1, T2 and T3.

**Fig 8.**
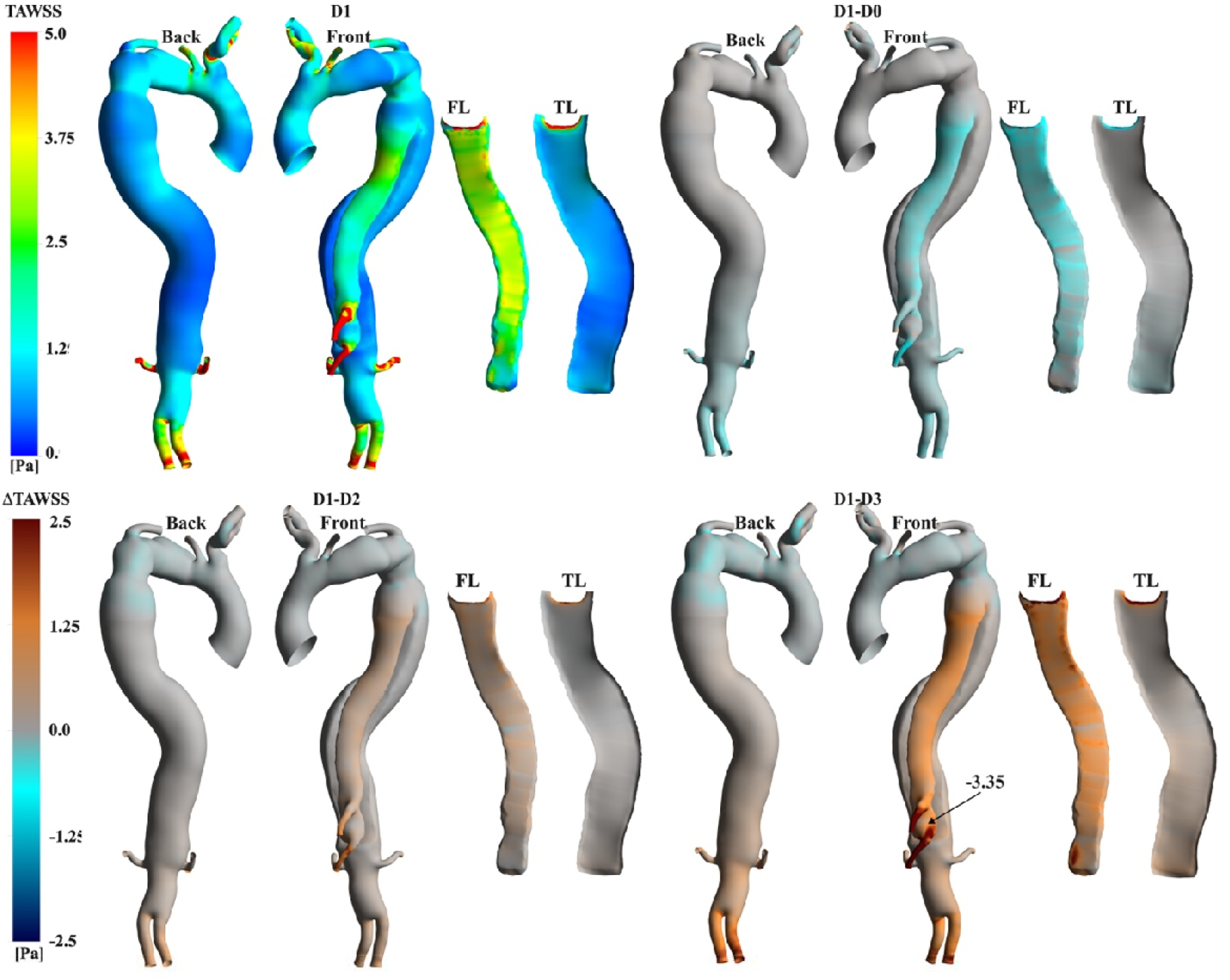
Contours of TAWSS for the patient-specific simulation (D1) and point-wise differences against additional cases.

Figure 8 displays qualitatively similar TAWSS distributions. High values (>5 Pa) are found at the outlets and PET, where high velocities occur. Significant point-wise differences are observed between the cases at the TL, celiac trunk (CT), and superior mesenteric artery (SMA) locations. TAWSS values tend to be lower for D0 at the FL and higher for more mobile IF simulations, with the highest difference being -3.35 Pa at the SMA compared to D3.

The highly fluctuating OSI indicates disturbed flow in both lumina in all cases (Figure 9). Notably, regions of high OSI are present at the arch, along both lumina and proximal to the VAO. The D1-D0 point-wise differences highlight that the rigid IF simulation slightly underpredicts OSI values at the FL. Conversely, higher OSI values are predicted at the TL in D2 and D3. Additionally, proximal to the VAO at the bottom of the FL, where higher displacements occur, a -0.36 point-wise difference is measured in the D1-D3 comparison (Figure 9).

**Fig 9.**
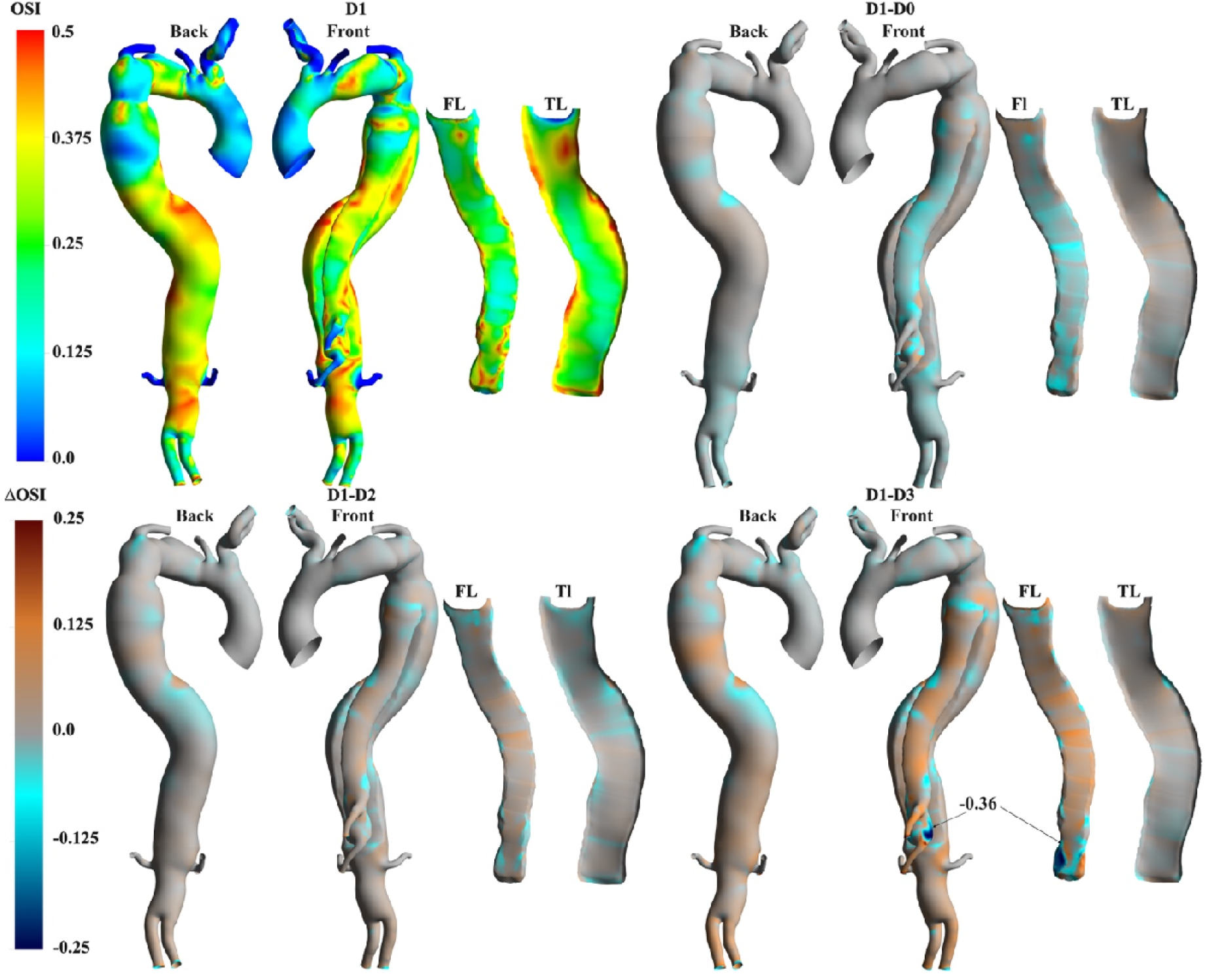
Contours of OSI for D1 and point-wise differences against additional cases

Relatively low TAWSS and fluctuating OSI lead to high RRT (>25 Pa^-1^) at both lumina and the VAO in D1 (Figure 10). Specifically, the highest RRT observed is 910.55 Pa^-1^ at the DA. The rigid flap simulation does not capture this localised region of elevated RRT. Conversely, this high RRT region is accentuated in D2 and D3, with the maximum point-wise difference reaching -1649.43 Pa^-1^ in D3.

**Fig 10.**
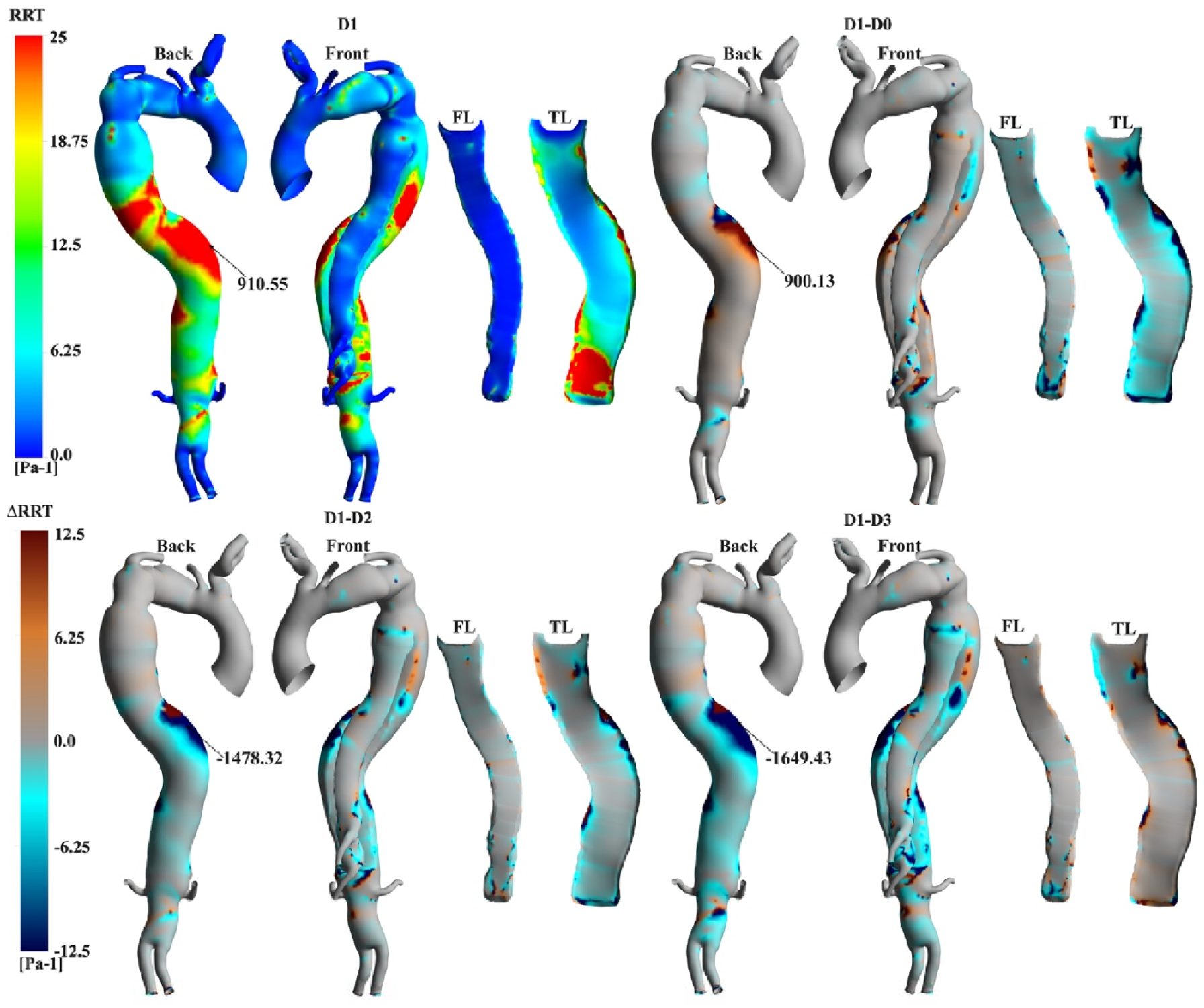
Contours of RRT for D1 and point-wise differences against additional cases.

## 3. Discussion

This study aimed to understand the impact of IF displacement in TBAD hemodynamics by enhancing a previously developed MBM and using exclusively 4DMRI and brachial pressures as input clinical data.

Quantifying IF mobility, a highly patient-specific variable, has been explored in various studies, revealing its crucial role in influencing malperfusion, haemodynamics, and treatment efficacy throughout different stages of TBAD [40], [41]. IF displacement is typically found to range from 0 to 3mm in chronic cases. The patient-specific case studied here (D1) revealed detailed insights into the interplay between intimal flap (IF) displacement, transmural pressure (TMP), and associated area changes throughout the cardiac cycle. These findings align with literature observations, demonstrating that these factors are associated with TL compression, FL expansion, and increased risk of rupture [42], [43]. At peak systole (T1), the IF displacement reaches its maximum, resulting in TL compression and FL expansion due to high pressure within the FL (Figure 4-5). During the deceleration phase, these FL/TL-compression/expansion patterns were inverted as the IF moved back towards its diastolic position, promoting a positive TMP and aligning with the reduction of cross-sectional area at the VAO (Figure 6) and neglecting the motion of IF in numerical simulations of TBAD (D0) results in underpredicted and low TMP at every time point in the cardiac cycle. This suggests that rigid IF simulations may lead to inaccurate luminal remodelling predictions, as a TMP close to zero is associated with better FL remodelling outcomes [9], [44].

On the other hand, simulations assuming a more mobile IF exhibited higher pressures within the FL during peak systole. This facilitated higher cross-sectional area variations, with the most significant expansions noted at the VAO for the D3 case. This suggests that a more mobile IF can exacerbate higher TMP, potentially influencing luminal expansion and promoting disturbed flow conditions. The TMP trends observed align with previous studies (Figure 4), which proposed a correlation between IF motion and disease progression [22], [26]. This highlights the potential of our model to further explore the mechanism of disease progression, particularly for highly mobile IF.

Compliant models, whether experimental or simulation-based, have been shown to more accurately mimic aortic flow compared to 4DMRI, according to recent research [25], [45], [46]. Studies on TBAD showed that 4DMRI provides a good qualitative overview of flow patterns in regions of interest within the aorta. However, 4DMRI fails to accurately quantify flows in low-velocity and highly aneurysmal regions, where a poor signal-to-noise ratio (SNR) degrades flow measurement quality [47], [48]. Conversely, CFD simulations informed by 4DMRI, as in the present study, offer an opportunity for reliable haemodynamic analyses [31], [49] even in regions that 4DMRI fails to capture, such as, for example, flow velocity regions during diastole and near wall hemodynamics (Figure 2-8,10). The predicted flow patterns in the patient-specific simulations (D1) agreed well with 4DMRI, especially proximal to IF during systole, where the high velocities of the true lumen (TL), linked with high pressure and IF displacement, were well captured. The rigid IF simulation (D0) showed underpredicted TL and overpredicted FL velocity magnitudes, as depicted at T1 and T2 at the PET, DA and VAO (Figure 2), despite comparing well with 4DMRI in certain locations, such as distal to the IF at the AA, due to the similar boundary conditions employed. Higher flow rate differences were observed with a more mobile IF than in vivo measurements, with errors reaching up to 4.5% at the visceral branches. These results provide key insights into the biomechanics of malperfusion in which a highly mobile IF can alter flow through visceral branches.

The complex geometry of the aortic arch has been found to induce rotational flow and heightened vorticity, potentially contributing to the development of aortic dissection [50], [51]. Furthermore, the presence of rotational flow structures has been recently linked to the remodelling and growth of localised regions in both TBAD and reconstructed TBAD [31], [52]. Naim et al., 2016, found that vortical structures dominated the FL in a TBAD study. They added that these structures expanded and clustered around the entry tear during systole, causing frequent platelet collisions and likely promoting thrombus formation. Additionally, recent studies demonstrated that the intensity and the topology of helical flow structures can be affected when comparing compliant to rigid wall simulations [54], [55]. In our study, the vorticity contours at the PET in D1 highlighted the presence of counter-rotating vortices, leading to high IRF values (Figure 7). The most complex patterns were observed at peak systole (T1). Such vortical patterns were less evident at the DA and VAO, where a separation between clockwise and anticlockwise vorticity was clear at both lumina. Swirling flows are not well captured when a rigid IF was assumed (D0) in regions where high IF displacements were predicted, for example, at the VAO during systole (T0, T1 and T2). Similar locations of vorticity with higher magnitudes were simulated when the IF reached higher displacements in D2 and D3 (Figure 7). Swirling structures observed with the streamlines had different shapes at peak systole, such as at the PET and VAO, demonstrating that the magnitude of the IF displacement has a noticeable impact on the WSS [56].

The IRF, which gives a measure of the intensity of the rotation of the flow on a plane, has been found to promote aortic and FL growth and dilation when it is high [37], [57], [58]. A similar study also indicated that high IRF can be linked to low WSS and, hence, the promotion of thrombosis (Ruiz-Muñoz et al., 2024). Some estimated IRF values along the lumina in our patient-specific simulation (D1) can be considered high, as indicated in the literature, as they were >20 cm^2^/s (Figure 7). The highest IRF values correspond to the vortical structures observed at the PET and VAO at peak systole. The patient-specific 3DIVP used as an inlet boundary condition promoted high circulation intensity at the AA at T1 and T2. In contrast, the IF motion appeared to have no impact, as evidenced by the negligible differences in IRF values between the patient-specific case (D1) and other simulation cases. However, discrepancies in IRF values can be noted at the PET and VAO when comparing D0 with D1, indicating the shortcoming of rigid IF assumption in fully capturing the rotational nature of the flow. The increased IF mobility in D2 and D3 led to higher IRF magnitudes and a shift in flow direction within the false lumen (FL) at T1. These IRF values are induced by flow pattern changes, which can affect areas of stagnation and recirculation, impacting WSS distribution in the FL even after TEVAR. Such alterations in WSS may influence the risk of further vessel wall damage and patient outcomes.

Research has shown that IF motion impacts flow dynamics and pressures and plays a crucial role in thrombus formation dynamics [24]. Abnormal aortic haemodynamics have been shown to affect WSS distributions and associated metrics [13]. For example, colocation of low TAWSS and high OSI has been linked to aortic growth, thrombosis and high RRT [60], [61]. Thus, high OSI values, triggered by the flow circulation close to the visceral branches, and low TAWSS in the distal and narrowed portion of the FL in D1 (Figure 9) suggest the likelihood of cell deposition therein. Such conditions were also observed at the DA. More specifically, RRT >900 Pa^-1^ values were predicted at the DA, coinciding with locations of chaotic flow and indicating a high potential for aortic growth (Figure 10). Similar observations can be made at the PET, where a high TAWSS>5Pa, due to a high velocity and chaotic vortical structures forming at the luminal flow separation location, could promote a risk of aneurysmal formation or local wall rupture (Figure 8). D0 failed to simulate this pattern close to the SMA, emphasising the importance of compliant IF simulation for remodelling predictions in TBAD.

The MBM employed in this study makes certain simplifications, such as assuming a linear relationship between displacement and force. Due to limitations in the temporal resolution of 4DMRI and cine-MRI, obtaining a transient description of the discrete radial and non-elastic behaviour of the aorta was impossible. Additionally, since only one plane of cine-MRI captures the displacement of the IF, a constant stiffness had to be applied across the entire IF. Although the wall and intimal flap measurements taken from the cine-MRI fall within the resolution error margin of the imaging technique, the simulation results remained consistent with these measurements, demonstrating the accuracy of the model. The method assumes that the pair of IF patches share the same normal, singular coordinate system, which should be considered for every node. This would be computationally heavy, and since the comparison of D1 against in vivo data showed good accuracy, the approach proposed here was deemed an acceptable compromise. Further research will aim to gather in vivo data with a finer resolution for a patient cohort. However, the insights gained from this study can still provide valuable information for understanding the hemodynamic effects of IF mobility in similar cases.

In conclusion, this study provided insights into the critical impact of IF mobility on TBAD haemodynamics using an improved MBM informed by 4DMRI data. By accurately simulating the patient-specific IF displacement, models revealed that increased IF mobility exacerbated TMP and promoted disturbed flow conditions, potentially leading to luminal expansion, thrombus formation, and aortic rupture. The findings underscored the importance of using compliant IF models over rigid simulations for accurate remodelling predictions and the impact of IRF, vorticity and WSS on disease progression and treatment outcomes. Clinically, these insights could inform more effective intervention strategies, such as tailored surgical planning to mitigate the adverse effects of a mobile IF and better understand whether or not a conservative approach might lead to optimal outcomes in terms of the natural resolution of TBAD. Future research should focus on gathering higher-resolution *in vivo* data to refine these models further and enhance their clinical applicability.

## Data Availability

All data produced in the present study are available upon reasonable request to the authors

## Acknowledgements

We thank the Department of Mechanical Engineering at University College London, the Wellcome EPSRC Centre for Interventional Surgical Sciences (WEISS) (203145/A/16/Z), the British Heart Foundation (NH/20/1/34705), the Biotechnology and Biological Sciences Research Council (BBSRC) and UK Research and Innovation (UKRI) (BB/X005062/1) for their funding support.

We gratefully acknowledge the support from the Computer Science Department at University College London and access to their high-performance computing facilities.

