## Supplemental materials for "Impact of residual intimal flap displacement post-TEVAR on TBAD haemodynamics in compliant, patient-specific CFD simulations informed by MRI"

### 1. Mesh Sensitivity Analysis

In this research, key haemodynamic metrics were analyzed across three refined meshes using the Grid Convergence Index (GCI) for validation purposes [1]. The meshes were created using Fluent Mesh (Ansys Inc., USA), employing a size field based on curvature and proximity, with a growth rate of 1.2 and the maximum and minimum cell sizes listed in Table 1. To accurately represent the turbulent boundary layer, ten prism layers were used, with the initial layer thickness calibrated to achieve a y+ value of approximately 1. A medium mesh, referred to as M2, with cell sizes ranging from 2 mm to 0.5 mm, was part of a mesh independence study. M2 was compared against a coarser mesh, M1, and a finer mesh, M3. M1 and M3 were created by doubling and halving the cell sizes, respectively. The element count ratio between M1 and M2, as well as between M2 and M3, is about 50%. Detailed element count data for each mesh is provided in Table 1:

**Table 1** Element count of M1, M2 and M3 and percentage change between M1/M2 and M2/M3.

| | **Mesh** | | | **Change** | |
|---|---|---|---|---|---|
| | M1 | M2 | M3 | M1/M2 | M2/M3 |
| **Element Count** | 924994 | 1931786 | 4251967 | 47.9% | 45.4% |
| **Minimum Element Size[mm]** | 1 | 0.5 | 0.25 | | |
| **Maximum Element Size [mm** | 4 | 2 | 1 | | |

The mesh quality and analysis were evaluated across seven planes and regions of interest (refer to Figure 1). Mean and maximum velocities were measured on the planes, while the mean time-averaged wall shear stress (TAWSS) was recorded in each region. The relative error between the metrics was calculated for both M1 versus M2 and M2 versus M3. The GCI was computed as follows:

$$r \sim \left(\frac{N_3}{N_2}\right)^{1/3} \sim \left(\frac{N_3}{N_2}\right)^{1/3} \quad (1)$$

$$p = \frac{\ln\left(\frac{|f_1-f_2|}{|f_2-f_3|}\right.}{\ln(r)} \quad (2)$$

$$E_{2,1}=\frac{|f_1-f_2|}{f_2.(r^p-1)} \quad (3)$$

$$E_{3,2}=\frac{|f_2-f_3|}{f_3.(r^p-1)} \quad (4)$$

$$GCI_{2,1}=F_s|E_2| \quad (5)$$

$$GCI_{3,2}=F_s|E_3|$$
(6)

With $f_{1,2,3}$ the metric of interest for M1, M2 and M3, $N_{1,2,3}$ denote the number of elements in M1, M2 and M3 respectively. $f_{1,2,3}$ is the evaluated metric for each mesh. The safety factor, $F_S$, was set at 1.25 and was defined by Celik et al., [2] and utilised by Armour et al.,[3].

The relative error in the metrics of interest between M2 and M3 was less than 4.6%. Additionally, $GCI_{3,2}$ did not exceed 4.3%, consistent previous study [4]. Consequently, the medium mesh, M2, was selected for the study and subsequent analysis.

| Mean TAWSS | | | | | | | |
|---|---|---|---|---|---|---|---|
| **Region** | **A** | **B** | **C** | **D** | **E** | **F** | **G** |
| **M1 [Pa]** | 1.229 | 1.026 | 0.566 | 0.238 | 1.268 | 0.771 | 1.049 |
| **M2 [Pa]** | 1.200 | 1.009 | 0.565 | 0.236 | 1.370 | 0.911 | 1.040 |
| **M3 [Pa]** | 1.2087 | 0.996 | 0.569 | 0.240 | 1.333 | 0.921 | 1.034 |
| **%M2,M1** | -2.4% | -1.7% | -0.1% | -1.0% | 8.1% | 18.2% | -0.8% |
| **%M3,M2** | 0.7% | -1.3% | 0.6% | 1.6% | -2.7% | 1.1% | -0.5% |
| **GCI 2,1** | 0.3% | 0.0% | 0.4% | 0.5% | 1.5% | 1.7% | 0.0% |
| **GCI 3,2** | 0.1% | 4.3% | 0.5% | 0.2% | 0.3% | 0.0% | 0.0% |

| Mean velocity | | | | | | | |
|---|---|---|---|---|---|---|---|
| **Plane** | **1** | **2** | **3** | **4** | **5** | **6** | **7** |
| **M1 [m/s]** | 0.1559 | 0.1632 | 0.0938 | 0.057 | 0.0881 | 0.0356 | 0.0356 |
| **M2 [m/s]** | 0.155 | 0.167 | 0.093 | 0.057 | 0.091 | 0.036 | 0.036 |
| **M3 [m/s]** | 0.1573 | 0.1746 | 0.0943 | 0.0588 | 0.0920 | 0.0353 | 0.0353 |
| **%M2,M1** | -0.3% | 2.3% | -0.5% | 0.5% | 2.7% | -0.3% | -0.3% |
| **%M3,M2** | 1.2% | 4.6% | 1.1% | 2.6% | 1.7% | -0.6% | -0.6% |
| **GCI 2,1** | 1.8% | 9.0% | 5.1% | 4.8% | 6.0% | 3.7% | 0.6% |
| **GCI 3,2** | 3.0% | 1.4% | 0.1% | 0.3% | 0.3% | 1.6% | 2.3% |

| Max Velocity | | | | | | | |
|---|---|---|---|---|---|---|---|
| **Plane** | **1** | **2** | **3** | **4** | **5** | **6** | **7** |
| **M1 [m/s]** | 0.5078 | 0.5737 | 0.2909 | 0.1367 | 0.3372 | 0.1318 | 0.1318 |
| **M2 [m/s]** | 0.503 | 0.571 | 0.289 | 0.136 | 0.336 | 0.132 | 0.132 |
| **M3 [m/s]** | 0.504 | 0.573 | 0.289 | 0.135 | 0.337 | 0.133 | 0.133 |
| **%M2,M1** | -0.9% | -0.4% | -0.6% | -0.9% | -0.3% | -0.2% | -0.2% |
| **%M3,M2** | 0.1% | 0.3% | 0.0% | -0.3% | 0.1% | 1.1% | 1.1% |
| **GCI 2,1** | 1.8% | 9.0% | 5.1% | 4.8% | 6.0% | 3.7% | 0.6% |
| **GCI 3,2** | 3.0% | 1.4% | 0.1% | 0.3% | 0.3% | 1.6% | 2.3% |

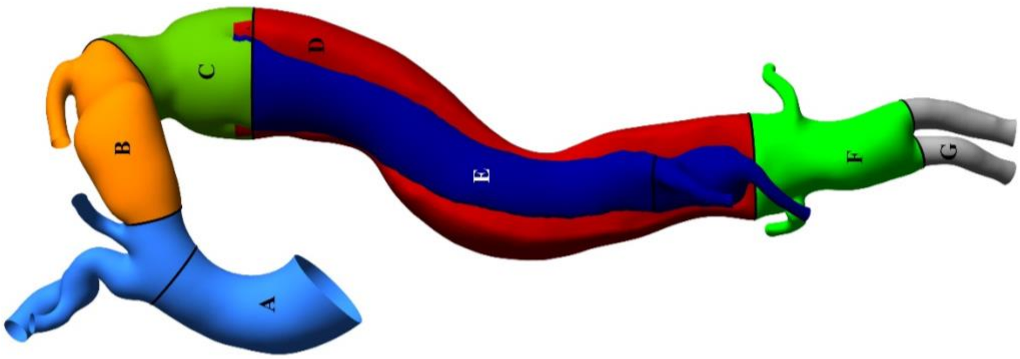


**Figure 1** Mean TAWSS, mean and max velocity relative error and GCI comparisons. Below the tables is the geometry on which regions and planes of interest, denoted by capital letetrs and numbers, respectively, are depicted.

### 2. Three Element Windkessel Parameters

Below is a table grouping the three-element Windkessel parameters used at the outlets of the aorta to apply a pressure condition mimicking the effects of the peripheral vasculature system [5]. The abbreviations stand for right common carotid (RCC), right subclavian (RSA), left common carotic (LCC), left subclavian (LSA), coeliac trunk (CT), superior mesenteric (SMA), right renal (RR), left renal (LR), left iliac (LI) and right iliac (RI). Considering the compliance of the aorta downstream of the arch in S2 after the specific stiffness leads to an increase in aortic compliance.

**Table 1** Three-element Windkessel parameters used for all simulatations in the paper.

| | R2[mmhg/mL.s] | R1[mmhg/mL.s] | C[mL/mmHg |
|---|---|---|---|
| **RCC** | 9.03 | 0.54 | 0.16 |
| **RSA** | 13.05 | 0.77 | 0.11 |
| **LCC** | 13.82 | 0.82 | 0.11 |
| **LSA** | 8.69 | 0.52 | 0.17 |
| **CT** | 26.58 | 0.77 | 0.06 |
| **SMA** | 15.91 | 0.46 | 0.09 |
| **LR** | 10.08 | 3.92 | 0.11 |
| **RR** | 8.16 | 3.17 | 0.13 |
| **LI** | 14.91 | 0.43 | 0.10 |
| **RI** | 16.82 | 0.48 | 0.09 |
